# Ketamine and attentional bias to threat: dynamic causal modeling of magnetoencephalographic connectivity in treatment-resistant depression

**DOI:** 10.1101/2021.02.22.21252247

**Authors:** Jessica R. Gilbert, Christina S. Galiano, Allison C. Nugent, Carlos A. Zarate

**Affiliations:** Experimental Therapeutics and Pathophysiology Branch, National Institute of Mental Health, National Institutes of Health, Bethesda, Maryland, USA

## Abstract

The glutamatergic modulator ketamine rapidly reduces depressive symptoms in individuals with treatment-resistant major depressive disorder (MDD) and bipolar disorder. While its underlying mechanism of antidepressant action is not fully understood, modulating glutamatergically-mediated connectivity appears to be a critical component moderating antidepressant response. This double-blind, crossover, placebo-controlled study analyzed data from 19 drug-free individuals with MDD and 15 healthy volunteers who received a single intravenous infusion of ketamine hydrochloride (0.5 mg/kg) as well as an intravenous infusion of saline placebo. Magnetoencephalographic recordings were collected prior to the first infusion and six to nine hours after both drug and placebo infusions. During scanning, participants completed an attentional dot probe task that included emotional faces. Antidepressant response was measured across timepoints using the Montgomery-Asberg Depression Rating Scale (MADRS). Dynamic causal modeling (DCM) was used to measure changes in parameter estimates of connectivity via a biophysical model that included realistic local neuronal architecture and receptor channel signaling, modeling connectivity between the early visual cortex, fusiform cortex, amygdala, and inferior frontal gyrus. Clinically, ketamine administration significantly reduced depressive symptoms in MDD participants. Within the model, ketamine administration led to faster gamma aminobutyric acid (GABA) and N-methyl-D-aspartate (NMDA) transmission in the early visual cortex, faster NMDA transmission in the fusiform cortex, and slower NMDA transmission in the amygdala. Ketamine administration also led to direct and indirect changes in local inhibition in the early visual cortex and inferior frontal gyrus and to indirect increases in cortical excitability within the amygdala. Finally, reductions in depressive symptoms in MDD participants post-ketamine were associated with faster α-amino-3-hydroxy-5-methyl-4-isoxazolepropionic acid (AMPA) transmission and increases in gain control of spiny stellate cells in the early visual cortex. These findings provide additional support for the GABA and NMDA inhibition and disinhibition hypotheses of depression and support the role of AMPA throughput in ketamine’s antidepressant effects.

## 1. Introduction

Ketamine’s rapid antidepressant effects have galvanized research into the neurobiological underpinnings of mood disorders and have increased focus on the potential role that the glutamatergic and GABAergic systems play in the etiology and pathophysiology of both major depressive disorder (MDD) (Bernard et al., 2011; Choudary et al., 2005; Luscher et al., 2011) and bipolar depression (Eastwood and Harrison, 2010). As a result of promising clinical and preclinical data, interest in investigating the glutamate system has grown exponentially (Ohgi et al., 2015), with many studies focusing on ketamine and its glutamatergically-modulating metabolites as viable clinical treatment options (Diazgranados et al., 2010; Zanos et al., 2019; Zarate et al., 2006). A wealth of studies have now demonstrated that a single infusion of subanesthetic-dose ketamine can rapidly (within hours) relieve depressive symptoms in individuals with both MDD (Murrough et al., 2013; Zarate et al., 2006) and bipolar depression (Diazgranados et al., 2010; Zarate et al., 2012), including those who are treatment-resistant. Repeat-dose studies have also pointed to continued improvements over longer time periods compared with a single administration (Phillips et al., 2019). Understanding the mechanism of action underlying ketamine’s rapid antidepressant effects could help identify novel biomarkers of antidepressant response and expedite the development of novel, rapid-acting therapeutics capable of more effectively treating depressive symptoms without the psychotomimetic side effects and risk for misuse associated with ketamine.

Ketamine is a noncompetitive N-methyl-D-aspartate (NMDA) receptor antagonist. Nevertheless, a host of studies suggest the possibility that NMDA receptor antagonism may not be the direct mechanism underlying ketamine’s antidepressant effects, and several other mechanisms are being investigated. For instance, recent studies found that the ketamine metabolite (2R,6R)-hydroxynorketamine (HNK) exerts antidepressant effects in animal models even though it is not an NMDA receptor antagonist at therapeutically relevant concentrations (Lumsden et al., 2019); rather, (2*R*,6*R*)-HNK appears to exert antidepressant effects by enhancing α-amino-3-hydroxy-5-methyl-4-isoxazolepropionic acid (AMPA) throughput (Zanos et al., 2016).

In addition, subanesthetic-dose ketamine administration leads to immediate disinhibition of glutamatergic neurons, producing a glutamate surge (Moghaddam et al., 1997). This surge is thought to result from NMDA receptor blockade by ketamine of fast-spiking gamma-aminobutyric acid (GABA)-ergic interneurons, leading to local inhibition of interneuron tonic firing and the subsequent disinhibition of pyramidal neurons (Duman et al., 2019; Homayoun and Moghaddam, 2007). Due to NMDA receptor blockade on post-synaptic excitatory neurons, excess synaptic glutamate is primarily taken up by AMPA receptors, thereby activating neuroplasticity-related signaling pathways, including mammalian target of rapamycin complex 1 (mTORC1) (Li et al., 2010; Li et al., 2011) and brain-derived neurotrophic factor (BDNF) (Liu et al., 2012), both of which result in increased synaptic potentiation and synaptogenesis. Furthermore, a host of cascading intracellular changes following ketamine administration involve eukaryotic elongation factor 2, which promotes BDNF release (Autry et al., 2011; Monteggia et al., 2013) and homeostatic synaptic scaling mechanisms (Kavalali and Monteggia, 2020); cellular changes resulting from direct inhibition of extrasynaptic NMDA receptors (Miller et al., 2014) activate plasticity mechanisms and also promote synaptic potentiation.

Within the field of psychiatry, a growing body of evidence suggests that altering the ratio of cortical excitation/inhibition balance could underlie a host of disorders, including depression (Fogaça and Duman, 2019; Godfrey et al., 2018). Preclinical work has also demonstrated that therapeutic-dose ketamine reduces inhibitory input onto pyramidal cells, thereby increasing synaptically-driven pyramidal cell excitation in single cell and population-level electrophysiological recordings (Widman and McMahon, 2018). Modeling work has robustly demonstrated that gamma rhythms reflect a balance between network-level excitation and inhibition (Buzsáki and Wang, 2012; Economo and White, 2012; Ray and Maunsell, 2015). In addition, work from our laboratory and that of others found that therapeutic-dose ketamine administration leads to robust increases in gamma power (Cornwell et al., 2012; Muthukumaraswamy et al., 2015; Sanacora et al., 2014; Shaw et al., 2015), potentially reflecting alterations in excitation-inhibition balance associated with antidepressant response (Cornwell et al., 2012; Nugent et al., 2019a; Nugent et al., 2019b).

Emotional processing deficits have been extensively reported in MDD. For example, compared to healthy volunteers, individuals with MDD showed a bias towards negative emotional information (Dalgleish and Watts, 1990; Mathews and MacLeod, 1994), including a bias towards faces demonstrating negative emotions compared to positive emotions (Gotlib et al., 2004; Joormann and Gotlib, 2007). One task of particular interest is the dot probe attentional task, which has been used to study emotional biases in depression (Peckham et al., 2010). Several neuroimaging studies have identified activation differences between healthy volunteers and participants with MDD using a dot probe task (Amico et al., 2012; Hu et al., 2017; Reed et al., 2018); anxiolytic (Ironside et al., 2016) and pharmacological treatment effects (Reed et al., 2018) on task performance in depression have also been observed.

This study sought to model ketamine-mediated differences in brain network connectivity during an attentional dot probe task with emotional faces in a group of participants with treatment-resistant MDD and healthy volunteers who underwent both ketamine and placebo saline infusions. This double-blind, crossover, placebo-controlled study used magnetoencephalography (MEG) in tandem with dynamic causal modeling (DCM) to model effective connectivity at three timepoints: a) baseline, b) six to nine hours following subanesthetic (0.5 mg/kg) ketamine infusion, and c) six to nine hours following placebo saline infusion. DCM uses a biophysical model that includes realistic local neuronal architecture to model effective connectivity between regions of interest (ROIs). Model inversion—the fitting of parameterized mean-field neuronal models to electrophysiological data features—results in *in silico* parameter estimates that govern unobservable neuronal states such as receptor-mediated connectivity between cell populations (here, a lumped estimate of AMPA/NMDA and GABA for excitatory and inhibitory intrinsic connections, respectively, in addition to AMPA and NMDA drive estimates for all extrinsic connections) and decay times of specific receptor types (here, AMPA, GABA, and NMDA) (Moran et al., 2011b). DCM was used to estimate connectivity in a fully reciprocally connected network of regions activated by the task, including the early visual cortex, fusiform cortex, amygdala, and inferior frontal gyrus. Because the study focused on measuring parameters that were significantly altered following ketamine administration, the post-ketamine scan was directly compared with both the baseline and placebo saline scans. It was predicted that ketamine would increase gamma power in our defined network—particularly in the amygdala—in line with previous findings of gamma power as a putative marker of ketamine-mediated synaptic potentiation (Gilbert and Zarate, 2020) and a normalizer of activation in the amygdala post-ketamine administration (Reed et al., 2018). The study also sought to examine group (MDD participants versus healthy volunteers) by session (ketamine versus baseline/placebo) interaction effects on modeled parameter estimates governing receptor time constants and connectivity within the amygdala, a key region involved in the emotional processing of face stimuli.

## 2. Material and Methods

### 2.1 Participants

All participants were studied at the National Institute of Mental Health (NIMH) in Bethesda, Maryland between September 2011 and August 2016. The present study used data drawn from a larger clinical trial (NCT00088699) that assessed ketamine’s antidepressant effects. The present study comprised 19 individuals with a DSM-IV-TR diagnosis of MDD (American Psychiatric Association, 1994) without psychotic features (11 F, mean age=36.7±10.9 years) and 15 healthy volunteers (11F, mean age=34.7±11.8 years). This subset of participants was selected because they had usable MEG scans for all three sessions of interest. Individuals with MDD were 18 to 65 years old, were experiencing a major depressive episode lasting at least four weeks, had not responded to at least one adequate antidepressant trial during the current major depressive episode, and had a Montgomery-Asberg Depression Rating Scale (MADRS) (Montgomery and Asberg, 1979) score of ≥ 20 at screening. Diagnosis was determined by Structured Clinical Interviews for Axis I DSM-IV-TR Disorders (SCID)–Patient Edition (First et al., 2002). Healthy volunteers were also 18 to 65 years old, had no Axis I disorder as determined by the Structured Clinical Interviews for Axis I DSM-IV-TR Disorders – Non-Patient Edition, and had no family history of Axis I disorders in first-degree relatives. All MDD participants were hospitalized for the duration of the study and were drug-free from psychotropic medications for at least two weeks prior to MEG testing (five weeks for fluoxetine, three weeks for aripiprazole). Healthy volunteers completed study procedures as inpatients but were otherwise outpatients. All participants were also in good health as evaluated by a medical history and physical examination, toxicology screens and urinalysis, blood laboratory results, clinical MRI, and electrocardiogram. The Combined Neuroscience Institutional Review Board at the National Institutes of Health approved the study. All participants provided informed written consent and were matched with an NIMH advocate from the Human Subjects Protection Unit to monitor consent and participation.

### 2.2 Clinical measurements

The primary clinical outcome measure for MDD patients—the MADRS (Montgomery and Asberg, 1979)—was administered 60 minutes prior to infusions (both ketamine and placebo) and at multiple time points (230 minutes and Days 1, 2, and 3) following infusions. Clinical outcome for MDD participants was modeled using all available data, controlling for both the period-specific baseline (−60-minute rating of that infusion) as well as a participant-average baseline (averaging both −60-minute ratings) and infusion. Repeated observations were accounted for by freely estimating the residual variance and covariance for each participant/infusion by drug (i.e., unstructured covariance matrix estimated by drug). The difference between ketamine and placebo was then estimated at 230 minutes, the time point closest to the MEG scan.

### 2.3 MEG acquisition and preprocessing

MEG recordings were collected at baseline and six to nine hours following both ketamine and placebo saline experimenter-blinded infusions. Ketamine and placebo infusions occurred 14 days apart, with infusion order randomized across participants. During each session, participants completed a dot probe task with emotional face stimuli presented using E-Prime presentation software (Psychology Software Tools, Pittsburgh, PA). The task used a mixed block/event-related design. During each trial, a fixation cross was presented centrally for 500 msec, where the participant was instructed to maintain focus. This was followed by the presentation of two simultaneous, side-by-side faces for 500 msec. One face displayed a happy, angry, or neutral expression, while the other was always neutral. After each pair of faces, a single dot was presented for 200 msec behind one of the two faces, and participants were instructed to press a button to indicate the presentation side (left or right). Trials where the dot replaced the emotional face were considered congruent trials, as the expectation was that attention would be biased towards the emotional face. Trials where the dot replaced the neutral face were considered incongruent. Trials were randomized and counterbalanced for emotion, gender of face, side of emotional face, and side of probe. Each trial was followed by a 1300 msec blank interstimulus interval. Jitter was also randomly added to reduce expectancy effects, during which a central fixation cross was presented. Trials were additionally blocked into two “angry blocks” and two “happy blocks”, with block order randomized across participants. Angry blocks comprised trials with angry and neutral faces or two neutral faces. Happy blocks comprised trials with happy and neutral faces or two neutral faces. This resulted in four emotional face trial types: angry congruent, angry incongruent, happy congruent, and happy incongruent, each having 48 trials over the experimental run. In addition, because neutral pairs were included in both happy and angry blocks, there were a total of 96 neutral paired trials.

Neuromagnetic data were collected using a 275-channel CTF system with SQUID-based axial gradiometers (VSM MedTech Ltd., Couquitlam, BC, Canada) housed in a magnetically-shielded room (Vacuumschmelze, Germany). Data were collected at 600 Hz with a bandwidth of 0-300 Hz. Synthetic third order balancing was used for active noise cancellation. Offline, MEG data were first visually inspected, and trials were removed where visible artifacts (e.g., head movements, jaw clenches, eye blinks, and muscle movements) were present. Second, individual channels showing excessive sensor noise were marked as bad and removed from the analysis. Data were then bandpass filtered from 1-58 Hz and epoched from −100 to 1000 msec peristimulus time. The analysis routines available in the academic freeware SPM12 (Wellcome Trust Centre for Neuroimaging, London, UK, http://www.fil.ion.ucl.ac.uk/spm/) were used for data processing. This work used the computational resources of the NIH HPC Biowulf cluster (http://hpc.nih.gov).

### 2.4 Source localization and source activity extraction

The multiple sparse priors routine implemented in SPM12 was used to identify gamma frequency (30-58 Hz) sources of activity from each participant’s sensor-level data over a peristimulus event time window from −100 to 1000 msec. Gamma frequency was targeted, as recent findings in both animals and humans have demonstrated robust, ketamine-mediated cortical responses in that band (Cornwell et al., 2012; Hong et al., 2010; Lazarewicz et al., 2010; Muthukumaraswamy et al., 2015; Shaw et al., 2015), in keeping with ketamine’s ability to alter excitation-inhibition balance (Gilbert and Zarate, 2020). Induced responses to face pairs were localized to 512 potential mesh points using a variational Bayesian approach following co-registration of sensor positions to a canonical template brain. Participant-level activation maps were constructed following inversion of each session (i.e., baseline, placebo, ketamine) separately for all participants. No prior constraints on source location were used. Following the inversion, statistical maps of group activity were computed and a mixed-effects ANOVA was used to define source-localized cortical regions showing a main effect of task across all trial types, thresholded at p<0.05 family-wise error correction. Secondarily, the main effect of infusion (here, ketamine compared with placebo) was tested using a more liberal criterion of p<0.05, uncorrected.

Group-level statistical activation maps demonstrated stimulus-induced gamma-band activity in a network of brain regions including the bilateral early visual cortices, and extending into the parietal and frontal regions (Figure 1A). Because the study sought to characterize connectivity in a network of regions activated during visual processing of emotional faces, four regions were investigated in order to model forward and backward connections in a left-lateralized network: early visual cortex, fusiform cortex, amygdala, and inferior frontal gyrus (see Figure 1 and below for source locations). Early visual cortex, fusiform cortex, and inferior frontal gyrus were defined using their corresponding peak voxels from the average effect contrast in Figure 1A. Amygdala was defined using the peak voxel from the infusion contrast in Figure 1A. Subsequent DCM analyses focused on characterizing connectivity in these regions in a wide, 1-50 Hz frequency band to model stimulus-induced event-related potentials.

**Figure 1.**
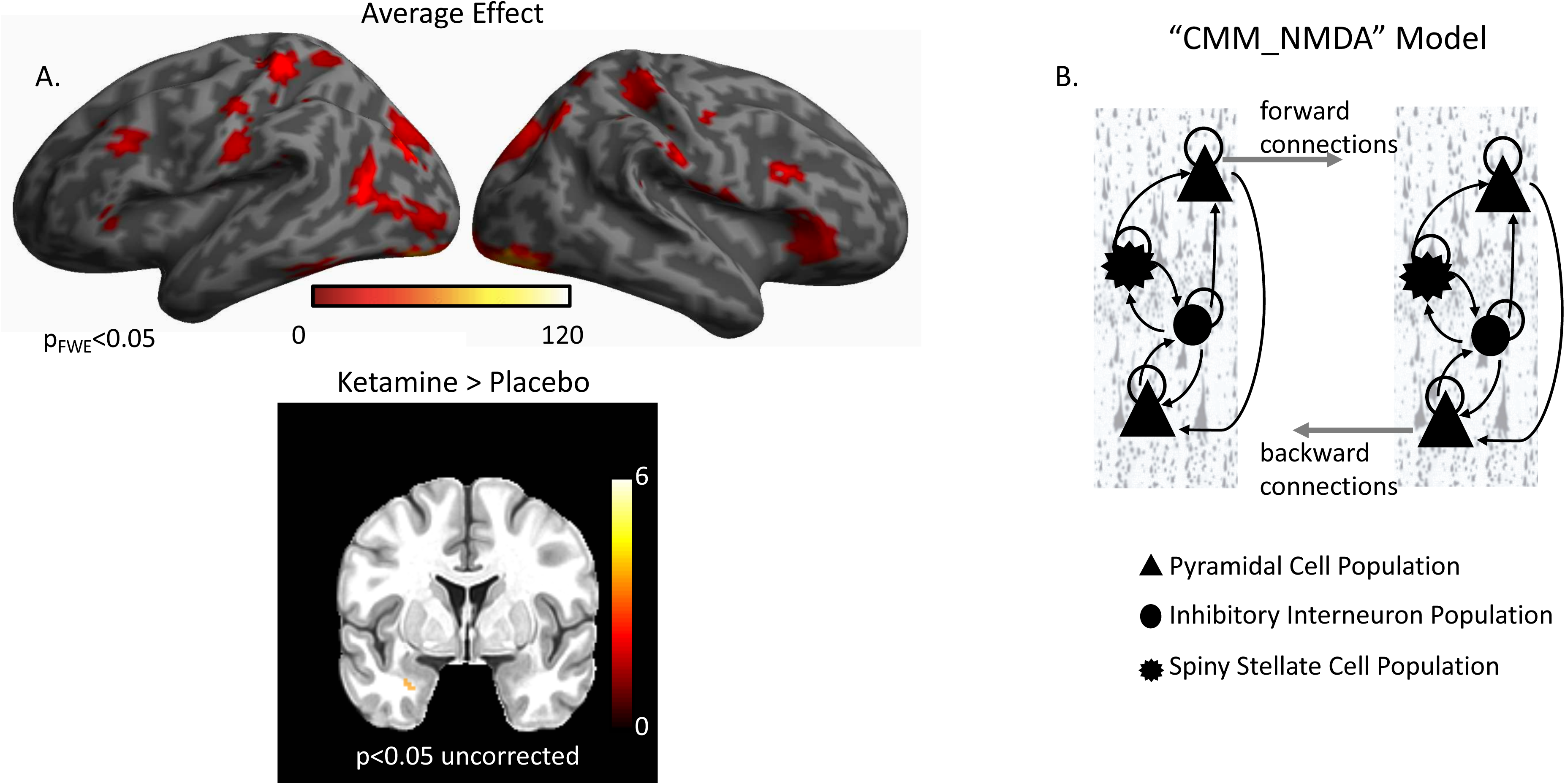
Gamma Power and Dynamic Causal Modeling (DCM). **(A)** A network of regions showed robust increases in induced gamma power during the task. These included the bilateral early visual cortex, bilateral fusiform cortex, and bilateral inferior frontal gyrus. When directly testing the effect of infusion, higher induced gamma power was found in the left amygdala for the ketamine infusion. **(B)** The default CMM_NMDA model includes four distinct intrinsic (within region) cell layers: superficial pyramidal cells, spiny stellate cells, inhibitory interneurons, and deep pyramidal cells. Intrinsic excitatory connections were mediated by α-amino-3-hydroxy-5-methyl-4-isoxazolepropionic acid (AMPA) and N-methyl-D-aspartate (NMDA) receptors while intrinsic inhibitory connections were mediated by gamma aminobutyric acid (GABA) receptors. Each cell population included a self-gain parameter that reflected precision for each cell type. Each receptor also included distinct time constants and dynamics in the model. Between regions, superficial pyramidal cells carry forward extrinsic signals to excitatory spiny stellate cells. Deep pyramidal cells carry backward extrinsic signals to both superficial pyramidal cells and inhibitory interneurons.

### 2.5 Dynamic causal modeling

DCM uses a biophysical model of neural responses based on neural mass models to predict recorded electrophysiological data (David et al., 2006). The present study specifically used the ‘CMM_NMDA’ model, a conductance-based neural mass model for electrophysiology, as implemented in SPM12 (http://www.fil.ion.ucl.ac.uk/spm/), to model responses between ROIs. The CMM_NMDA model includes connection parameters for AMPA- and NMDA-mediated glutamatergic signaling as well as GABA signaling. Within the model, superficial pyramidal cells encode and carry feedforward signaling to stellate cells, while deep pyramidal cells carry feedback signaling to superficial pyramidal cells and inhibitory interneurons (Figure 1B). Additional parameters include AMPA, GABA, and NMDA time constants, the inverse of which model the rate of receptor channel opening and closing within each ROI. The model has been extensively described in the literature, and detailed equations can be found elsewhere (Moran et al., 2011a; Muthukumaraswamy et al., 2015; Symmonds et al., 2018).

Thalamic (stimulus-bound) input was modeled with a Gaussian bump function that drove activity in early visual cortex (−8, −94, −8) in the model. Two models of message-passing were constructed between the early visual cortex, fusiform cortex (−52, −52, −22), amygdala (−25, −3, −16), and inferior frontal gyrus (−48, 28, −2) (see Figure 2A). The first model was a traditional bottom-up processing model that included forward connections from early visual cortex to fusiform cortex, fusiform cortex to amygdala, and amygdala to inferior frontal gyrus. Backward connections ensured reciprocal message-passing in a top-down hierarchy. Model 2 included two additional connections: direct forward and reciprocal backward connections between early visual cortex and inferior frontal gyrus. Face emotion modulated all region-to-region connections in both models (i.e., comparing trials in which happy versus angry faces appeared).

**Figure 2.**
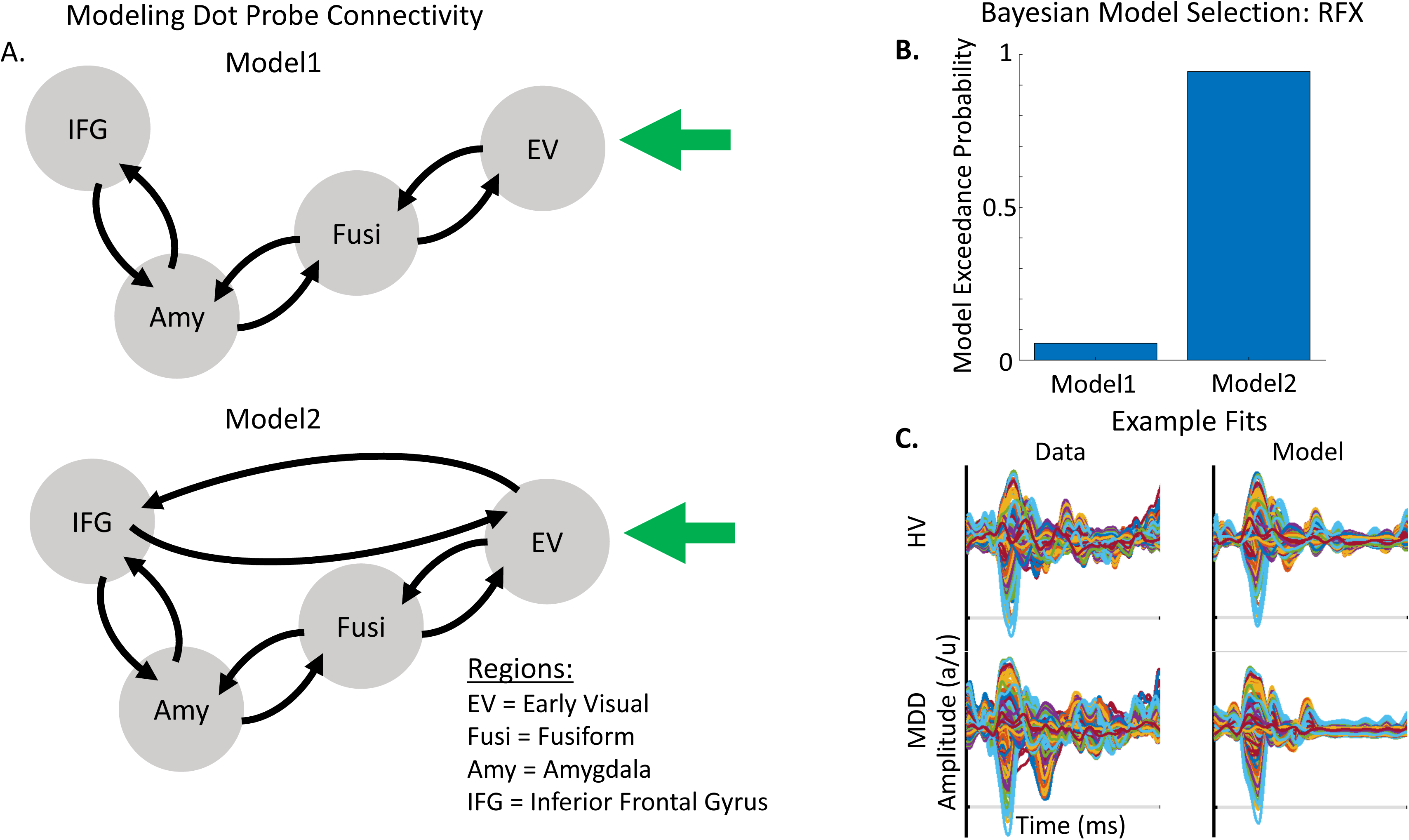
Models of Connectivity, Winning Model, and Example Model Fits. **(A)** Two plausible models were constructed to account for message passing between the early visual cortex (EV), fusiform cortex (Fusi), amygdala (Amy), and inferior frontal gyrus (IFG). Model 1 included reciprocal forward and backward connections from the EV to the Fusi, from the Fusi to the Amy, and from the Amy to the IFG. Model 2 included a direct, reciprocal, forward and backward connection from the EV to the IFG. **(B)** Bayesian Model Selection (BMS) was used to adjudicate between models, demonstrating that Model 2, with fully interconnected feedforward and feedback connections between each region, had the greatest exceedance probability. RFX: random effects. **(C)** Example evoked responses (left) and model fits (right) for a healthy volunteer (HV, top) and participant with major depressive disorder (MDD) (bottom).

For the DCM analyses, MEG activity for the extracted time series was fitted over 1-500 milliseconds peristimulus time in a wide frequency band from 1-50 Hz using an event-related potential (ERP) model to capture ERPs of evoked activity. For computational efficiency, DCM optimizes a posterior density over free parameters (parameterized by its mean and covariance) via a standard variational Bayesian inversion procedure (Friston et al., 2007). In the present analysis, initial DCMs were computed for each participant and session, and model fits were assessed. The posterior estimates were then used to initialize a second set of DCMs for each participant and session, and model fits were again assessed. This iterative procedure occurred for both Model 1 and Model 2. In both cases, the initialized model resulted in a better fit of the model to the data. The negative free energy bound on the log-model evidence was then used to adjudicate between Model 1 and Model 2 across participants, selecting the model with the greatest log-model evidence for subsequent analyses. Parameter estimates were extracted from optimized DCMs for the winning model for each participant and session to compare ketamine-mediated effects across parameter estimates.

To determine the mixture of parameters that mediated ketamine’s effects, a second-level modeling extension of DCM called parametric empirical Bayesian analysis (Friston et al., 2016) was applied. This analysis refits a full model (where all parameters can covary according to grouping) and provides reduced models where smaller combinations of parameters are considered and informed by differences between sessions. Group, session, and group by session effects on all parameters were specifically tested in the second-level design matrix, where the first column represented the average effect over all participants and sessions, the second column tested for the effect of group, the third column tested for the effect of drug, and the fourth column tested for group by drug interactions. Group by drug interactions were of particular interest, though group and drug effects are also reported here.

Finally, as additional exploratory analyses, post-hoc classical statistical tests were conducted to determine whether any parameters identified using parametric empirical Bayesian analysis as significantly contributing to group effect, drug effect, or group by drug interactions were associated with antidepressant response in the MDD participants only. Here, changes in parameter values from baseline to ketamine were specifically examined and correlated with changes in MADRS score from baseline to post-ketamine using pairwise linear correlation as implemented in MATLAB software. Because this analysis was exploratory, a liberal criterion of p<0.05, uncorrected, was used.

## 3. Results

Clinically, the effect of drug at 230 minutes post-ketamine infusion compared to 230 minutes post-placebo infusion was significant (t(18)=2.07, p<0.05), for an estimated reduction of 5.37 (SE=2.28) points on total MADRS score (95% CI: −0.05, +9.48) following ketamine administration. Behaviorally, both reaction time bias (calculated as the difference between congruent and incongruent trials for happy and angry faces, respectively) and accuracy rates on the emotional dot probe task were examined using multi-way ANOVAs to look for main effects of group, session (baseline, placebo, ketamine), emotion (happy versus angry), and congruency (congruent versus incongruent; calculated for accuracy scores only). In addition, all two-, three-, and four-way interactions were considered. Although no significant behavioral effects were observed on reaction time bias scores, main effects were found for group (F=14.43, p<0.01) and session (F=3.58, p<0.05) on accuracy scores for participants; in particular, MDD participants were more accurate (mean=94.2%) than healthy volunteers (mean=88.8%). In addition, both MDD participants and healthy volunteers were most accurate during the baseline session (mean=94.2%) followed by the ketamine session (mean=91.6%) and the placebo session (mean=89.8%). Post-hoc tests using Bonferroni correction found significant accuracy differences between the baseline and placebo sessions across participants (t=3.45, p<0.05).

MEG data were subsequently source-localized to infer the primary generators of the signal using the multiple sparse priors routine. Significant group-level induced gamma-band activation was identified in response to the dot probe task (Figure 1A). The network of regions activated included the bilateral early visual cortex extending into higher-order visual areas in the occipital lobe, regions of the temporal lobe including the fusiform gyrus, and regions in both the parietal and frontal lobes, including the inferior frontal gyrus. When testing for the effect of infusion (ketamine versus placebo), left-lateralized amygdala response was found at the more liberal criterion of p<0.05, uncorrected. We therefore focused on characterizing parameter estimates of effective connectivity using DCM for electrophysiology using a model that included left-lateralized early visual cortex, fusiform cortex, amygdala, and inferior frontal gyrus (Figure 2A).

Two plausible models were constructed to account for connectivity between ROIs. Using Bayesian model selection to adjudicate between these models, Model 2—which included the addition of forward and backward connections between the early visual cortex and inferior frontal gyrus—was found to have the strongest model evidence (Figure 2B). Example model fits for an MDD participant and a healthy volunteer are shown in Figure 2C.

Parametric empirical Bayes—an analysis approach that allows testing of random effects of model parameters at the group level—was used to test for parameters contributing to the group effect, drug effect, and group by drug interactions. All fitted parameters in the model were considered, focusing on parameters that exhibited meaningful effects (specifically, parameters having a probability of 95% or greater). All identified parameters are reported in Tables 1-3, and parameters showing meaningful group by drug interactions are reported here. Four receptor time constants showed meaningful group by drug interactions, including the GABA time constant in the early visual cortex and the NMDA time constants in the early visual cortex, fusiform cortex, and amygdala (Figure 3A). As the inverse of time constants are rate constants, faster rates of GABA and NMDA signal transmission were found in the early visual cortex for MDD participants post-ketamine, while healthy volunteers showed slower GABA signal transmission coupled with faster NMDA signal transmission following ketamine. In the fusiform cortex, faster NMDA signal transmission was observed for MDD participants post-ketamine, while healthy volunteers showed slower signal transmission. Finally, slower NMDA signal transmission in amygdala was observed for both groups post-ketamine.

**Figure 3.**
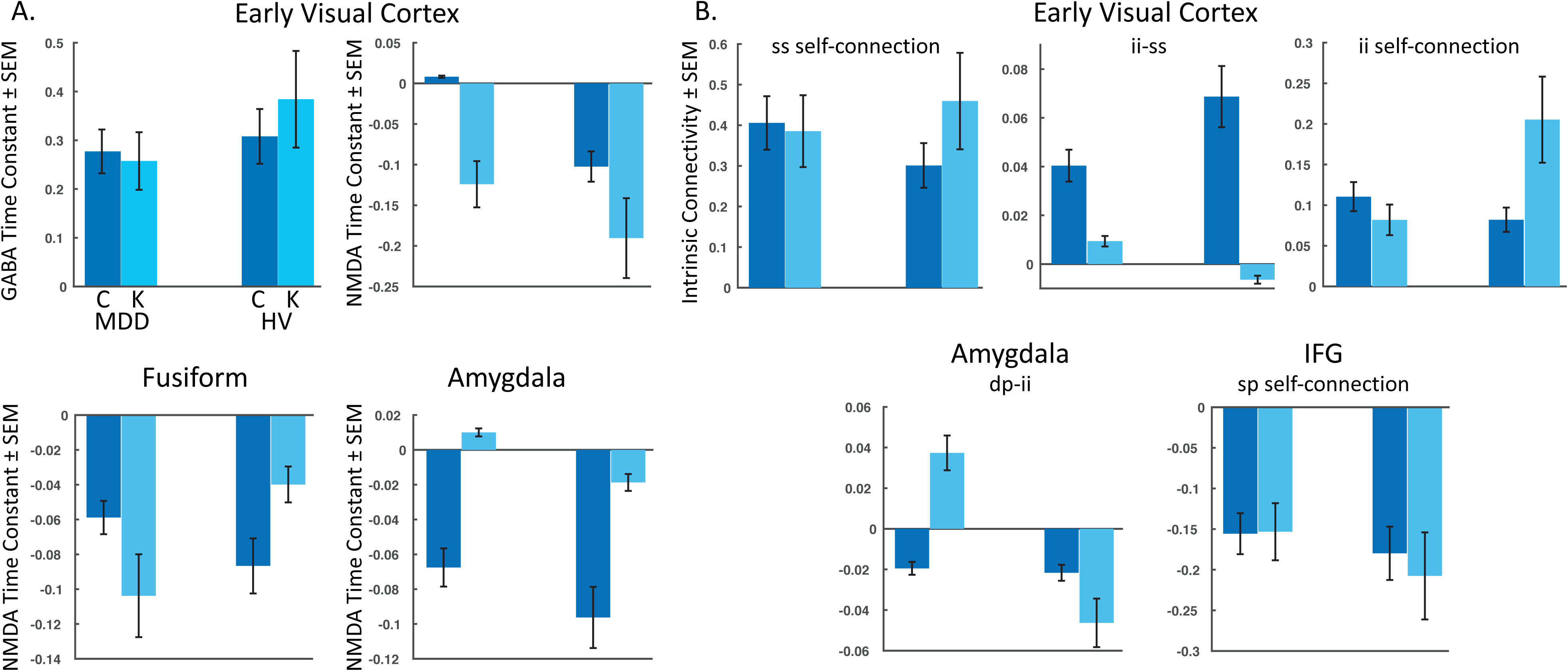
Meaningful Parameters Showing Group by Drug Interactions. The estimated log mean and variance of each meaningful (95% probability or greater) parameter are plotted for participants with major depressive disorder (MDD) (left) and healthy volunteers (HV) (right) for: **(A)** the four receptor time constants showing group by drug interactions, and **(B)** the five intrinsic connectivity parameters showing group by drug interactions. IFG=inferior frontal gyrus, C=baseline/placebo sessions, K=ketamine session, ss=spiny stellate cells, ii=inhibitory interneurons, sp=superficial pyramidal cells, dp=deep pyramidal cells.

**Table 1.** Group Effects Over Parameters

|  | Parameter | Parameter Estimate (Ep) | Posterior Probability (Pp) |
| --- | --- | --- | --- |
|  | <u>Time Constants</u> |  |  |
| 1 | AMPA - Amy* | -0.0978 | 1 |
| 2 | GABA – Fusi* | 0.1501 | 1 |
| 3 | GABA – IFG* | 0.1534 | 1 |
| 4 | NMDA – EV* | 0.1763 | 1 |
| 5 | NMDA – IFG* | -0.3113 | 1 |
|  | <u>Intrinsic Connectivity</u> |  |  |
| 6 | EV: inhibitory self-connection – ss* | 0.1285 | 1 |
| 7 | EV: inhibitory self-connection – ii | 0.0604 | 0.531 |
| 8 | Fusi: inhibitory self-connection – ss* | 0.2202 | 1 |
| 9 | Fusi: inhibitory self-connection – sp* | 0.1363 | 1 |
| 10 | Amy: excitatory connection – sp to dp* | -0.1424 | 1 |
| 11 | IFG: inhibitory self-connection – ss* | 0.1437 | 1 |

Note: Parametric empirical Bayes was used to identify the mixing of parameters that contributed to the effect of group. Note that the timing of data collection (six to nine hours post-ketamine administration) occurred past the half-life of ketamine. Meaningful parameters were defined as those with a posterior probability (Pp) greater than 95%. Ten parameters were found to significantly contribute to group effects. These included the α-amino-3-hydroxy-5-methyl-4-isoxazolepropionic acid (AMPA) time constant within the amygdala (Amy), gamma aminobutyric acid (GABA) time constants within the fusiform gyrus (Fusi) and inferior frontal gyrus (IFG), and N-methyl-D-aspartate (NMDA) time constants within the early visual cortex (EV) and IFG. In addition, the inhibitory self-connections on spiny stellate cells (ss) within the EV, Fusi, and IFG differed between groups, as did the inhibitory self-connection on superficial pyramidal cells (sp) within the Fusi, and the excitatory connections between sp and deep pyramidal cells (dp) in the Amy. Finally, the inhibitory self-connection on inhibitory interneurons (ii) in the EV showed a group effect, though not at our threshold. \*Pp>0.95.

**Table 2.** Drug Effects Over Parameters

|  | Parameter | Parameter Estimate (Ep) | Posterior Probability (Pp) |
| --- | --- | --- | --- |
|  | <u>Time Constants</u> |  |  |
| 1 | AMPA - EV* | 0.132 | 1 |
| 2 | GABA – Amy* | 0.1363 | 1 |
| 3 | GABA – IFG* | 0.1349 | 1 |
| 4 | NMDA – EV* | -0.2028 | 1 |
| 5 | NMDA – Amy* | 0.5303 | 1 |
| 6 | NMDA – IFG* | -0.2419 | 1 |
|  | <u>Intrinsic Connectivity</u> |  |  |
| 7 | EV: excitatory connection – sp to dp* | 0.1988 | 1 |
| 8 | EV: inhibitory connection – ii to sp* | -0.2011 | 1 |
| 9 | IFG: inhibitory self-connection – ss | 0.0773 | 0.502 |
| 10 | IFG: excitatory connection – ss to ii* | 0.1805 | 1 |

Note: Parametric empirical Bayes was used to identify the mixing of parameters that contributed to the effect of drug. Meaningful parameters were defined as those with a probability (Pp) greater than 95%. Nine parameters were found to significantly contribute to drug effects. These included the α-amino-3-hydroxy-5-methyl-4-isoxazolepropionic acid (AMPA) time constant within the early visual cortex (EV), gamma aminobutyric acid (GABA) time constants within the amygdala (Amy) and inferior frontal gyrus (IFG), and N-methyl-D-aspartate (NMDA) time constants within the EV, Amy, and IFG. In addition, the excitatory connections between superficial pyramidal cells (sp) and deep pyramidal cells (dp), and the inhibitory connections between inhibitory interneurons (ii) and sp differed following ketamine in the EV, as did excitatory connections between spiny stellate cells (ss) and ii in the IFG. Finally, the inhibitory self-connection on ss in the IFG showed a drug effect, though not at our threshold.
\*Pp>0.95.

**Table 3.** Group by Drug Interactions Over Parameters

|  | Parameter | Parameter Estimate (Ep) | Posterior Probability (Pp) |
| --- | --- | --- | --- |
|  | <u>Time Constants</u> |  |  |
| 1 | GABA - EV* | -0.1274 | 1 |
| 2 | NMDA – EV* | 0.1994 | 1 |
| 3 | NMDA – Fusi* | -0.1723 | 1 |
| 4 | NMDA – Amy* | 0.1502 | 1 |
|  | <u>Intrinsic Connectivity</u> |  |  |
| 5 | EV: inhibitory self-connection – ss* | -0.2778 | 1 |
| 6 | EV: inhibitory connection – ii to ss* | -0.1837 | 1 |
| 7 | EV: inhibitory self-connection – ii* | -0.4241 | 1 |
| 9 | Amy: excitatory connection – dp to ii* | 0.1998 | 1 |
| 10 | IFG: inhibitory self-connection – sp* | 0.1943 | 1 |

Note: Parametric empirical Bayes was used to identify the mixing of parameters that contributed to group by drug interactions. Meaningful parameters were defined as those with a probability (Pp) greater than 95%. Nine parameters were found to significantly contribute to group by drug effects. These included gamma aminobutyric acid (GABA) time constants within the early visual cortex (EV) and N-methyl-D-aspartate (NMDA) time constants within the EV, fusiform cortex (Fusi), and amygdala (Amy). In addition, the inhibitory self-connections on spiny stellate cells (ss) and inhibitory interneurons (ii), as well as inhibitory connections between ii and ss in the EV showed group by drug interactions. Excitatory connections between deep pyramidal cells (dp) and ii in the Amy, in addition to inhibitory self-connections on superficial pyramidal cells (sp) in the inferior frontal gyrus (IFG) also showed group by drug interactions. \*Pp>0.95.

Our second-level modeling extension also identified five intrinsic, within-region connections that showed meaningful group by drug interaction effects; three were in the early visual cortex, with one each in the amygdala and inferior frontal gyrus (Figure 3B). In the early visual cortex, decreased self-inhibitory drive was observed on both spiny stellate cells and inhibitory interneurons for MDD participants post-ketamine; in contrast, healthy volunteers showed increased self-inhibitory drive on both cell types post-ketamine. Ketamine was also found to reduce inhibitory drive from inhibitory interneurons to spiny stellate cells in the early visual cortex for both groups. In the amygdala, increased excitatory drive from deep pyramidal cells to inhibitory interneurons was noted for MDD participants post-ketamine, while healthy volunteers showed decreased excitatory drive between these connections. Finally, reduced self-inhibitory drive on superficial pyramidal cells in the inferior frontal gyrus was noted in healthy volunteers post-ketamine, but no changes were observed in MDD participants.

Finally, we explored whether any meaningful parameters identified as contributing to the group effect, drug effect, or group by drug interactions were associated with clinical change at 230 minutes post-ketamine compared to baseline. Two parameters were found to be associated with antidepressant response (Figure 4). First, change in AMPA time constants from baseline to ketamine were associated with antidepressant response in the MDD participants (r=0.4917, p<0.05), with faster AMPA signal transmission post-ketamine associated with better antidepressant response. Second, change in self-inhibitory drive of spiny stellate cells in early visual cortex from baseline to ketamine was associated with antidepressant response (r=-0.6545, p<0.01), with larger self-inhibition on spiny stellate cells post-ketamine associated with better antidepressant response.

**Figure 4.**
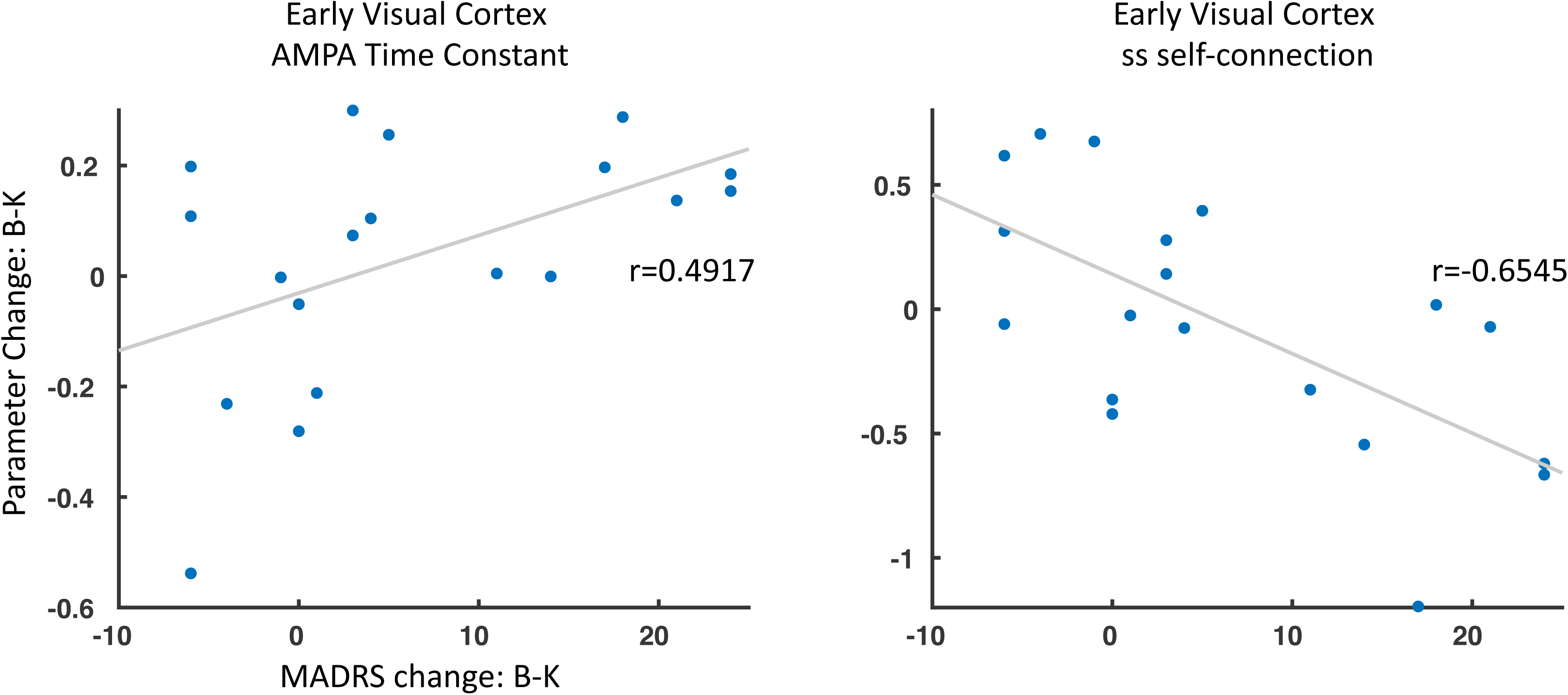
Parameter Change and Antidepressant Response. Two parameters showed a significant association between change from baseline (B) to ketamine (K) sessions and associated changes in Montgomery-Asberg Depression Rating Scale (MADRS) scores (B-K). ss=spiny stellate cells, AMPA= α-amino-3-hydroxy-5-methyl-4-isoxazolepropionic acid.

## 4. Discussion

This study used MEG recordings collected while participants completed a dot probe task with emotional faces in tandem with DCM to probe ketamine’s effects in individuals with treatment-resistant MDD and healthy volunteers. The goal was to measure *in silico* changes in connectivity within and between the early visual cortex, fusiform cortex, amygdala, and inferior frontal gyrus, in addition to changes in AMPA, GABA, and NMDA receptor time constants, following ketamine administration.

Clinically, we found significantly reduced depressive symptoms in our MDD sample post-ketamine, consistent with previous findings (Murrough et al., 2013; Zarate et al., 2006). Behaviorally, no differences in reaction time bias scores were observed on the task. However, accuracy differences were observed between the two groups, with MDD participants significantly more accurate than healthy volunteers during the task. In addition, session effects were noted with regard to accuracy rates, with the best performance occurring during the baseline session, followed by the ketamine and then placebo sessions. Importantly, post-hoc tests found significant differences in accuracy between the baseline and placebo sessions only. These findings suggest that healthy volunteers were less engaged in the task and therefore did not perform as well as the MDD participants. In addition, task repetition led to poorer performance, especially following placebo saline infusion, where participants were perhaps least motivated to perform well during the scan procedures.

We modeled induced gamma-band activity during the dot probe task, identifying a network of brain regions involved in the task. We also modeled regions showing an effect of infusion (ketamine versus placebo) and found increased gamma power in the amygdala post-ketamine versus placebo. These findings are in keeping with preclinical studies suggesting increased cortical excitation following ketamine administration, due to NMDA inhibition reducing the activity of putative GABA interneurons (Homayoun and Moghaddam, 2007). At a delayed rate, this increases the firing rate of pyramidal neurons due to enhanced AMPA throughput (Homayoun and Moghaddam, 2007) that, in turn, leads to increased cortical excitation. Given that gamma power in the amygdala showed a drug-specific effect, with increased cortical excitation post-ketamine, this suggests that increased cortical excitation in this key emotional face processing region may be related to normalization of emotional processing following drug administration. Notably, a similar normalization of amygdalar activity post-ketamine was previously described in an fMRI study that included an attentional dot probe task with emotional faces (Reed et al., 2018), though this was not specifically examined in the present study.

Two plausible models of message passing between the early visual cortex and the inferior frontal gyrus were subsequently fit. A model that included traditional feedforward processing along the ventral stream to the amygdala in tandem with feedforward connections from the early visual cortex to the inferior frontal gyrus provided the best model fits. All fitted parameters were subsequently extracted, and a Bayesian modeling extension of DCM was used to test for meaningful parameters contributing to the group effect, drug effect, and group by drug interactions. Here, we focus on discussing group by drug interactions, as these are identified parameters where ketamine had differential effects between MDD participants and healthy volunteers. Four receptor time constants showed group by drug interactions, including the GABA and NMDA time constants in the early visual cortex and the NMDA time constants in the fusiform cortex and amygdala. In the early visual cortex, ketamine administration led to faster GABA and NMDA transmission for MDD participants, while GABA transmission slowed for healthy volunteers post-ketamine. In the fusiform cortex, faster NMDA transmission followed ketamine administration for MDD participants, though the rate of transmission slowed for healthy volunteers post-ketamine. Interestingly, a slowing of NMDA transmission was observed in the amygdala post-ketamine for both MDD and healthy volunteers, though healthy volunteers had significantly faster NMDA transmission at baseline/placebo than MDD participants. As the amygdala ROI was identified based on the effect of infusion (ketamine versus placebo), slowing of NMDA transmission within this region is clearly related to drug effects. Although no association was noted between NMDA transmission in the amygdala and antidepressant response within our sample, future studies should examine whether these changes in NMDA time constants are related to other clinical measures of mood changes following drug administration.

In addition to changes in receptor time constants, group by drug interactions were found for intrinsic connectivity within the early visual cortex, amygdala, and inferior frontal gyrus. In the early visual cortex, three intrinsic connections showed group by drug changes in inhibitory drive. First, decreased GABAergic inhibitory drive on self-connections were found for both inhibitory interneurons and spiny stellate cells following ketamine in the MDD participants, while healthy volunteers demonstrated increased GABAergic inhibitory drive post-ketamine. These self-connections reflect gain or precision of different cell types, suggesting reductions in self-gain on inhibitory interneurons and spiny stellate cells following ketamine administration in the MDD group. Second, reduced inhibitory drive was observed on the intrinsic connection from inhibitory interneurons to spiny stellate cells in the early visual cortex in our MDD and healthy volunteers. Third, ketamine increased the excitatory drive from deep pyramidal cells to inhibitory interneurons in the amygdala in MDD participants, while healthy volunteers showed reduced excitatory drive for this connection post-ketamine. Finally, ketamine also reduced the inhibitory self-gain on superficial pyramidal cells in the inferior frontal gyrus in our healthy volunteers only. Interestingly, these findings all reflect changes in intrinsic connectivity that regulate or modulate inhibition locally. Within the amygdala in particular, increased excitatory drive onto inhibitory interneurons for MDD participants seems at odds with an increased state of excitability within this region; however, similar accounts of increased pyramidal-to-inhibitory interneuron drive have previously been reported (Shaw et al., 2020) and are thought to reflect a link between increased pyramidal cell excitability locally and downstream effects of increased gamma power.

Separately, we tested whether any meaningful parameters identified in our analysis of group effects, drug effects, or group by drug interactions were associated with antidepressant response in our MDD participants. We specifically examined changes in parameter estimates from the baseline to ketamine sessions (baseline minus ketamine) and correlated them with change in MADRS score from baseline to 230 minutes post-ketamine (the time point closest to the MEG recording session). Two parameters were found to be associated with antidepressant response, both in the early visual cortex. The first was the AMPA time constant in the early visual cortex, where faster AMPA transmission post-ketamine was associated with better antidepressant response. The second was inhibitory self-gain on spiny stellate cells in the early visual cortex, where larger self-inhibition on spiny stellate cells post-ketamine was associated with better antidepressant response. The findings of an association between AMPA transmission and antidepressant response are particularly striking because AMPA receptor throughput following NMDA receptor blockade (Duman et al., 2019; Moghaddam et al., 1997) is thought to result in delayed increases in synaptic potentiation and synaptogenesis, key mechanisms associated with ketamine’s antidepressant effects. Similar associations between AMPA receptor connectivity and antidepressant response were also previously reported in a time window overlapping with our MEG recordings (Gilbert et al., 2018; Gilbert et al., 2020).

One important limitation of this study is that MEG recordings were not collected during or immediately following infusions, but rather six to nine hours following ketamine administration in order to avoid side effects while measuring therapeutic drug effects. Thus, we cannot comment on acute changes in modeled parameter estimates. However, studies of ketamine’s acute effects in healthy volunteers suggest robust changes in both gamma power (Muthukumaraswamy et al., 2015; Shaw et al., 2015) and AMPA and NMDA receptor drive (Muthukumaraswamy et al., 2015) during ketamine infusion. Future studies should explore ketamine’s acute effects in MDD participants to better understand the mechanisms via which ketamine reduces depressive symptoms.

## 5. Conclusions

These findings demonstrate that ketamine administration leads to key changes in estimates of GABA and NMDA time constants measured using MEG in tandem with DCM. In addition to mirroring findings from animal studies measuring the acute effects of ketamine (Homayoun and Moghaddam, 2007), these changes also indicate that ketamine alters estimates of excitatory and inhibitory intrinsic connectivity within key regions important for visual processing of emotional faces. Finally, the findings also underscore the usefulness of DCM for modeling connectivity changes associated with ketamine administration.

## Data Availability

The data is available upon request from the corresponding author.

## Acknowledgements

The authors thank the 7SE research unit and staff for their support. Ioline Henter (NIMH) provided invaluable editorial assistance.

## Funding

This work was supported by the Intramural Research Program at the National Institute of Mental Health, National Institutes of Health (IRP-NIMH-NIH; ZIA MH002857), by a NARSAD Independent Investigator Award to Dr. Zarate, and by a Brain and Behavior Mood Disorders Research Award to Dr. Zarate. The funders had no further role in study design; in the collection, analysis, or interpretation of data; in the writing of the report; or in the decision to submit the paper for publication.

## Declaration of Interest

Dr. Zarate is listed as a co-inventor on a patent for the use of ketamine in major depression and suicidal ideation; as a co-inventor on a patent for the use of (2*R*,6*R*)-hydroxynorketamine, (*S*)-dehydronorketamine, and other stereoisomeric dehydro and hydroxylated metabolites of (*R,S*)-ketamine metabolites in the treatment of depression and neuropathic pain; and as a co-inventor on a patent application for the use of (2*R*,6*R*)-hydroxynorketamine and (2*S*,6*S*)-hydroxynorketamine in the treatment of depression, anxiety, anhedonia, suicidal ideation, and post-traumatic stress disorders. He has assigned his patent rights to the U.S. government but will share a percentage of any royalties that may be received by the government. All other authors have no conflict of interest to disclose, financial or otherwise.

## Notes

### Clinical Trial

NCT00088699

### Author Declarations

Combined Neuroscience Institutional Review Board at the National Institutes of Health

